# Tensor Fasciae Suralis – Prevalence Study and Literature Review

**DOI:** 10.1101/19010389

**Authors:** Logan S. Bale, Sean O. Herrin

## Abstract

Tensor fasciae suralis (TFS) is an accessory muscle of the posterior lower extremity. Although TFS has been documented in cadaveric and radiological reports, its prevalence remains unknown. The TFS variant is noteworthy to anatomists, as it may be encountered in the dissection laboratory, and clinicians, as the muscle could potentially cause confusion during physical examination or diagnostic imaging. Multiple variations of TFS have been reported in the literature, suggesting the need for a classification system. We dissected 236 formalin-fixed cadaveric lower limbs to determine the prevalence of TFS. The PubMed and MEDLINE databases were searched to compare the anatomical features of independent TFS case reports. In our prevalence study, the TFS muscle was identified in three lower limbs (1.3%). In total, 35 cases of TFS (31 cadaveric and four radiological) were identified in the literature. Our literature review revealed that the accessory muscle most often arises as a single head from the long head of the biceps femoris, yet many other presentations have been documented. The need for a classification system to distinguish between the subtypes of TFS became apparent during the literature review. Tensor fasciae suralis is a rare muscle, present in only 3 of 236 (1.3%) cadaveric lower limbs dissected in this study. We propose the use of a classification system, based on muscle origin and number of heads, to better organize the subtypes of TFS.

## INTRODUCTION

Tensor fasciae suralis (ischioaponeuroticus) is an accessory muscle of the posterior thigh and popliteal fossa. This supernumerary muscle can arise from various locations within the posterior thigh and inserts into the deep (sural, crural) fascia of the leg, gastrocnemius, or calcaneal tendon^1^. It has been suggested that TFS functions in leg flexion, although electrical studies have not been conducted to confirm this hypothesis^2^. As related by Schaeffer^3^, the first case of TFS was reported by Kelch^4^ and since then relatively few cadaveric^2,3,5–25^ reports of TFS have been published. We aimed to determine the prevalence of TFS by dissecting the posterior thighs and popliteal fossae of 236 cadaveric lower limbs. A literature review was conducted to assess the anatomical features of TFS that have been published in cadaveric and radiological case reports. A classification system that is focused on the muscle’s proximal attachment(s) is put forth herein to organize the subtypes of TFS. We also suggest that an often-used definition of TFS be modified and challenge the claim that the semitendinosus muscle is the most common point of origin of TFS. The clinical relevance of the accessory muscle is discussed.

## MATERIALS AND METHODS

### Prevalence Study

This study was exempt from IRB approval at University of Western States. As part of routine educational dissection, the lower limbs of 118 formalin-fixed cadavers (n = 236) were examined for the presence of the TFS muscle. The donor cadavers were of mixed demographics; there were 74 males (age range = 18-100 years) and 44 females (age range = 37-97 years).

### Literature Review

The following keywords were searched in the PubMed and MEDLINE databases: tensor fasciae suralis, ischioaponeuroticus, (variant OR variation OR anomaly AND posterior thigh OR popliteal fossa). Relevant publications were sought out in the reference lists of sources identified in the PubMed and MEDLINE searches. Muscle variants that arose from one or more of the hamstring muscles and inserted into the deep fascia of the leg, gastrocnemius, or calcaneal tendon were considered TFS. The authors have referenced original communications wherever possible, however secondary sources were used when primary sources could not be located. The following case reports were not included in the literature review: 1) a muscle that arose from the belly of semitendinosus and inserted into the peroneal retinaculum, peroneal trochlea of the calcaneus, and tuberosity of the fifth metatarsal that was determined to be the authors to be TFS^26^. We do not believe that this muscle variant was in fact TFS as its insertion points are not consistent with a muscle that could tense the sural fascia. 2) A different group related a case of TFS that was identified in a patient using MRI and stated that the same muscle variant was seen in an additional three patients^27^. Specific details related to the origin,insertion, and course of these four TFS cases were not explicitly discussed and as such they were excluded from the literature review. 3) the original communication (reported by Pardi^28^ and referenced by Barry and Bothroyd^23^) of a muscle that was likely TFS could not be located, nor could materials that discuss significant details of the case. Lastly 4), a unique supernumerary muscle of the popliteal fossa (termed femeroaponeuroticus by the authors) was excluded from this study as the muscle in question arose from the femur and was innervated by the common peroneal nerve^29^.

## RESULTS

### Prevalence Study

Of the 236 lower limbs included in this dataset, three TFS muscles were identified. One donor cadaver featured bilateral TFS muscles (Case 1). This case was previously reported by the authors of the current study^17^. A second donor cadaver featured the muscle variant in the left posterior thigh and popliteal fossa, but the right side was grossly unremarkable (Case 2).

#### Case 1 – 51-year-old male, height: 175 cm, weight: 68 kg

The TFS muscles identified in the posterior thighs and popliteal fossae of this cadaver were symmetrical. Each muscle arose from the long head of the biceps femoris by a fascial slip that measured approximately 3.3 cm in length (Fig. 1) and featured a fusiform muscle belly that was approximately 12.0 x 2.5 x 1.0 cm in greatest dimensions. Each TFS muscle inserted into the fascia of the medial and lateral heads of the gastrocnemius muscle.

**Figure 1:**
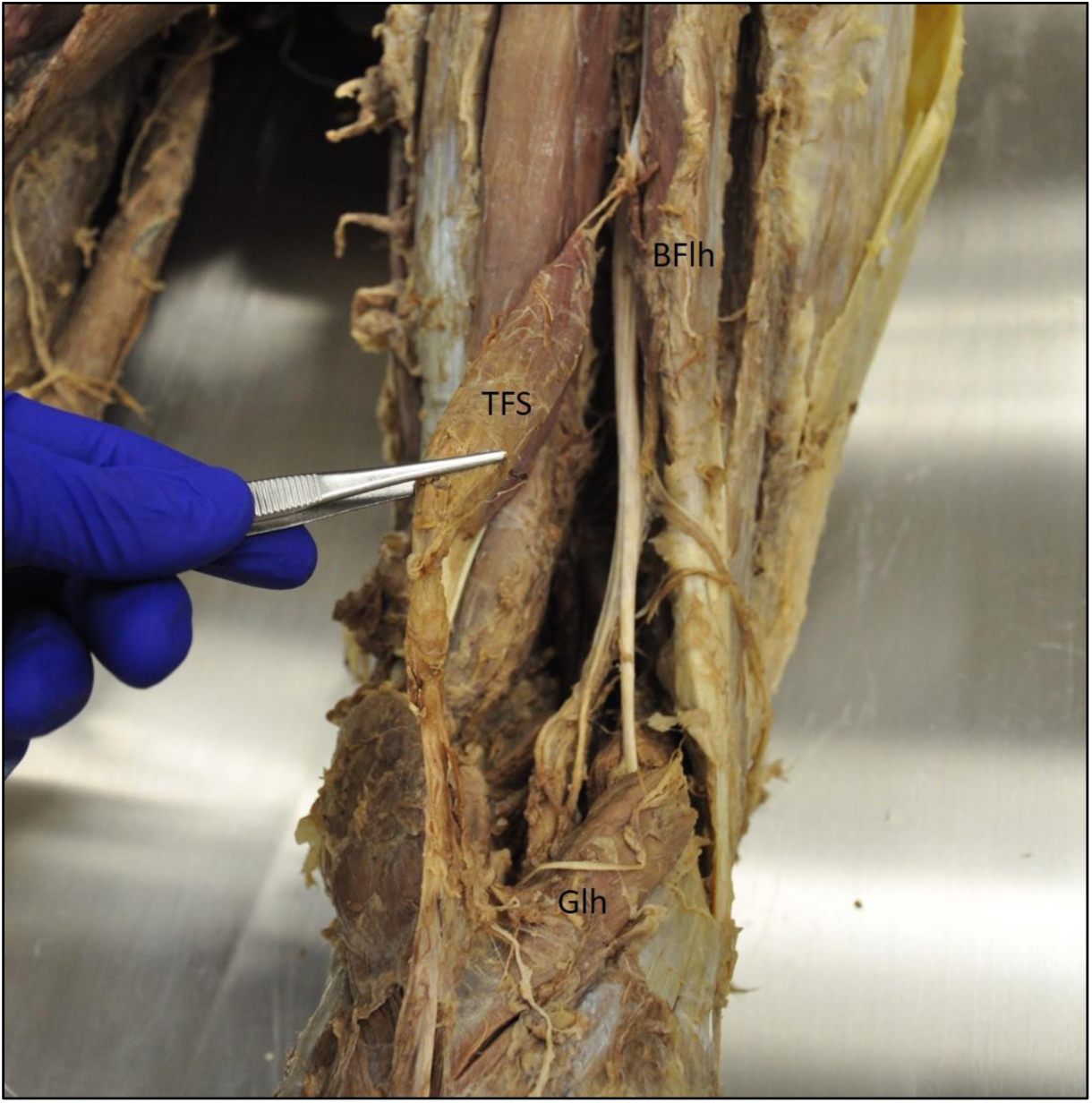
Right posterior thigh and popliteal fossa (**TFS**: tensor fasciae suralis muscle; **BFlh**: biceps femoris muscle, long head; **Glh**: gastrocnemius muscle, lateral head).

#### Case 2 – 68-year-old male, height: 178 cm, weight: 70 kg

The left posterior thigh and popliteal fossa contained a TFS muscle that arose from the long head of the biceps femoris approximately 18.5 cm distal to the common hamstring tendon (Fig. 2). The fusiform muscle belly was long and flat and measured approximately 22.0 x 1.0 x0.3 cm in greatest dimensions.The TFS muscle inserted into the sural fascia overlying the medial head of the gastrocnemius. The distal aspect of the muscle belly and fascial insertion of TFS were located immediately posterior to the medial sural cutaneous nerve.

**Figure 2:**
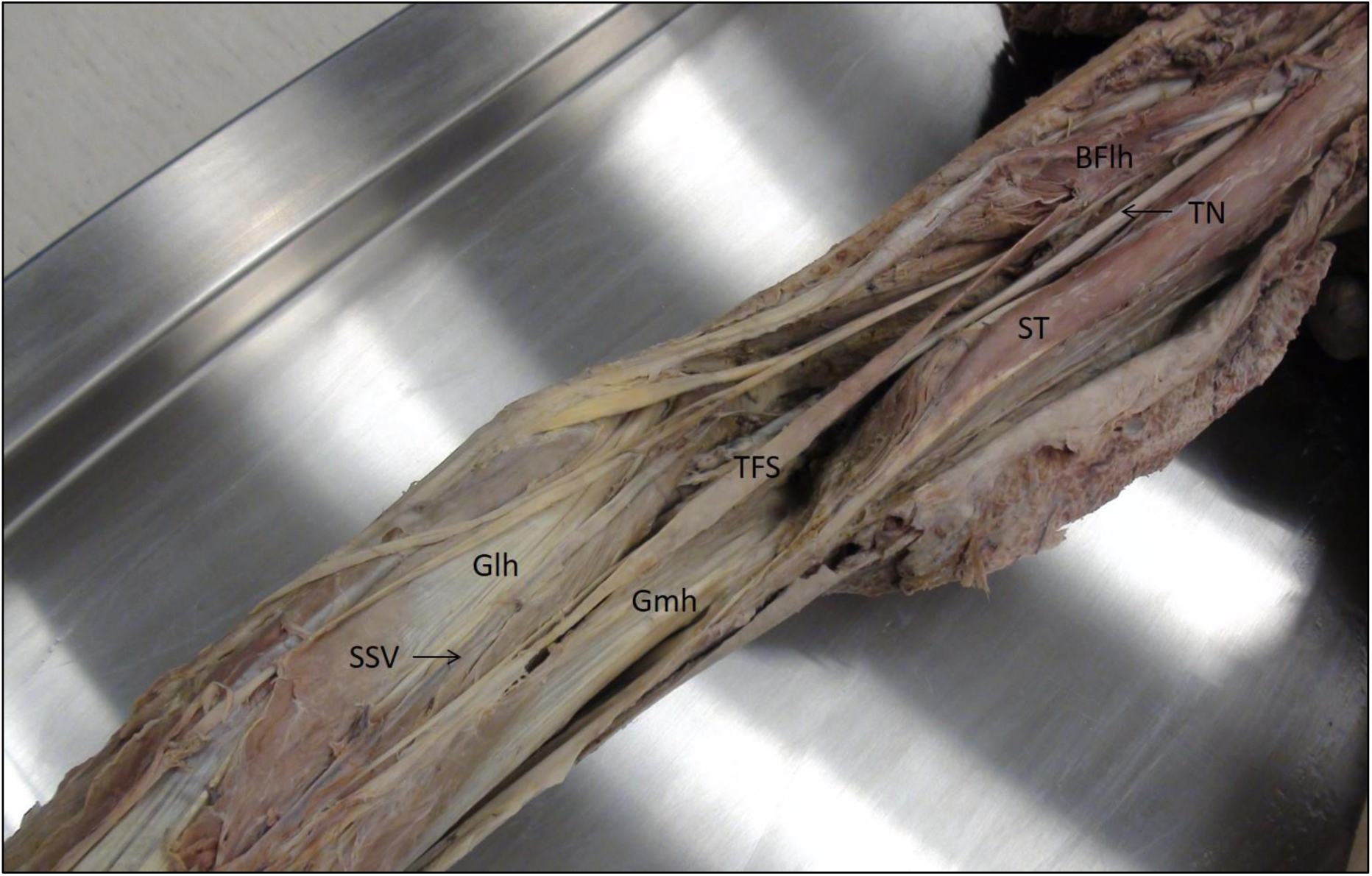
Left posterior thigh and popliteal fossa (**TFS:** tensor fasciae suralis muscle; **Glh:** gastrocnemius muscle, lateral head, **Gmh:** gastrocnemius muscle, medial head; **SSV:** small saphenous vein; **TN:** tibial nerve, **CFN:** common fibular nerve; **ST:** semitendinosus muscle, **SM:** semimembranosus muscle; **BFlh:** biceps femoris muscle, long head).

### Literature Review

A total of 31 cadaveric cases (25 unilateral, three bilateral) and four radiological cases (all unilateral) were identified in the literature. Unilateral cases were right-sided in 15 instances, left-sided in 10 instances, and not specified in four instances. Interestingly, the accessory muscle was found in only three female lower extremities compared to 25 male lower extremities (sex not specified in seven cases).

We propose a classification system (Table 1) to categorize the subtypes of TFS whereby the origin of the accessory muscle and number of heads present are used as distinguishing characteristics. The classification system is summarized below:

**Table 1:**
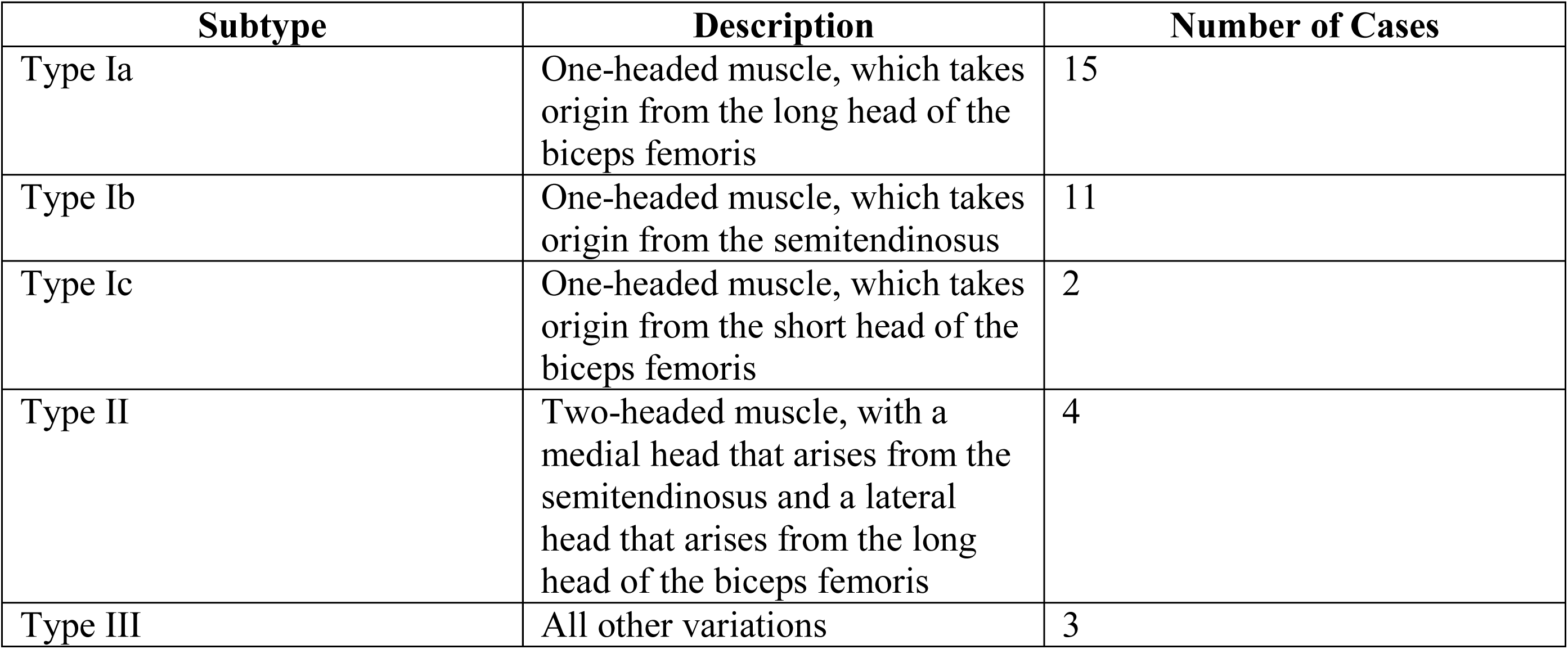
Overview of classification system for subtypes of TFS.

| Subtype | Description | Number of Cases |
| --- | --- | --- |
| Type Ia | One-headed muscle, which takes origin from the long head of the biceps femoris | 15 |
| Type Ib | One-headed muscle, which takes origin from the semitendinosus | 11 |
| Type Ic | One-headed muscle, which takes origin from the short head of the biceps femoris | 2 |
| Type II | Two-headed muscle, with a medial head that arises from the semitendinosus and a lateral head that arises from the long head of the biceps femoris | 4 |
| Type III | All other variations | 3 |

*Type I* – a one-headed TFS muscle which takes origin from the a) long head of biceps femoris, b) semitendinosus, or c) short head of biceps femoris.

*Type II* – a two-headed TFS muscle, with a medial head from semitendinosus and a lateral head from the long head of biceps femoris.

*Type III* – a TFS muscle that does not fit the criteria of the Type I or Type II subtypes.

Cases determined to be TFS are summarized in Table 2. When TFS arises as a single head (28 instances), it most commonly does so from the long head of the biceps femoris (15 instances), followed by the semitendinosus (11 instances), and short-head of the biceps femoris (two instances). When TFS arises as two heads (six instances), it typically does so from the long head of biceps femoris and semitendinosus (four instances), but other variations have been reported^21,23^. The accessory muscle arose from three heads in one case; each head took origin from a different hamstring muscle^9^. The most common insertion of TFS was the sural fascia, followed by the gastrocnemius, and lastly the calcaneal tendon. Several authors have reported that TFS is innervated by the tibial nerve^2,3,7–11,13,14,16,23,25^. The muscle’s blood supply has been noted to be from a branch of the femoral artery^23^ or a branch of the 4^th^ perforating artery of the profunda femoris^10^. We did not investigate the neurovascular supply of the TFS muscles that were identified in the current study.

**Table 2:**
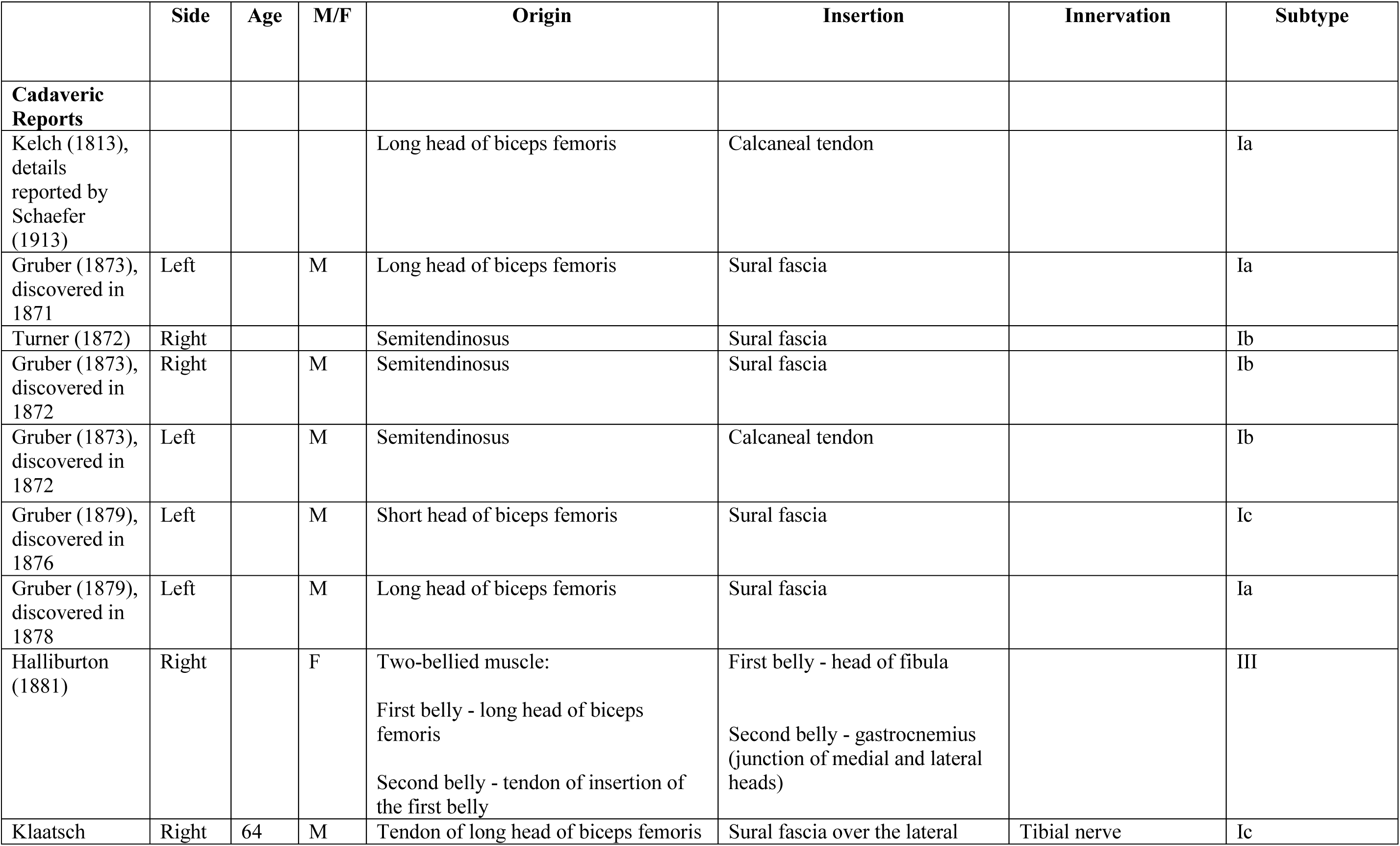
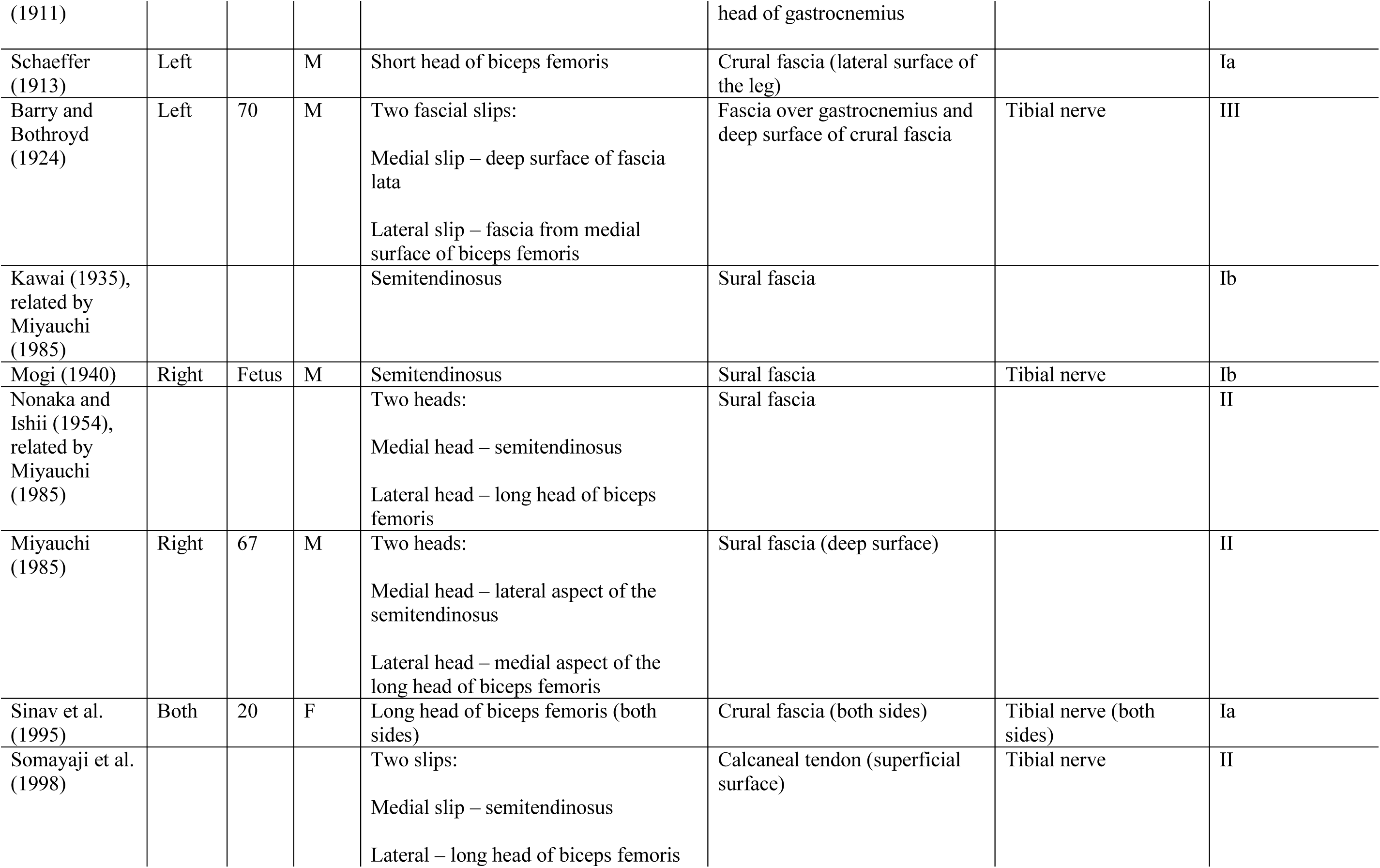
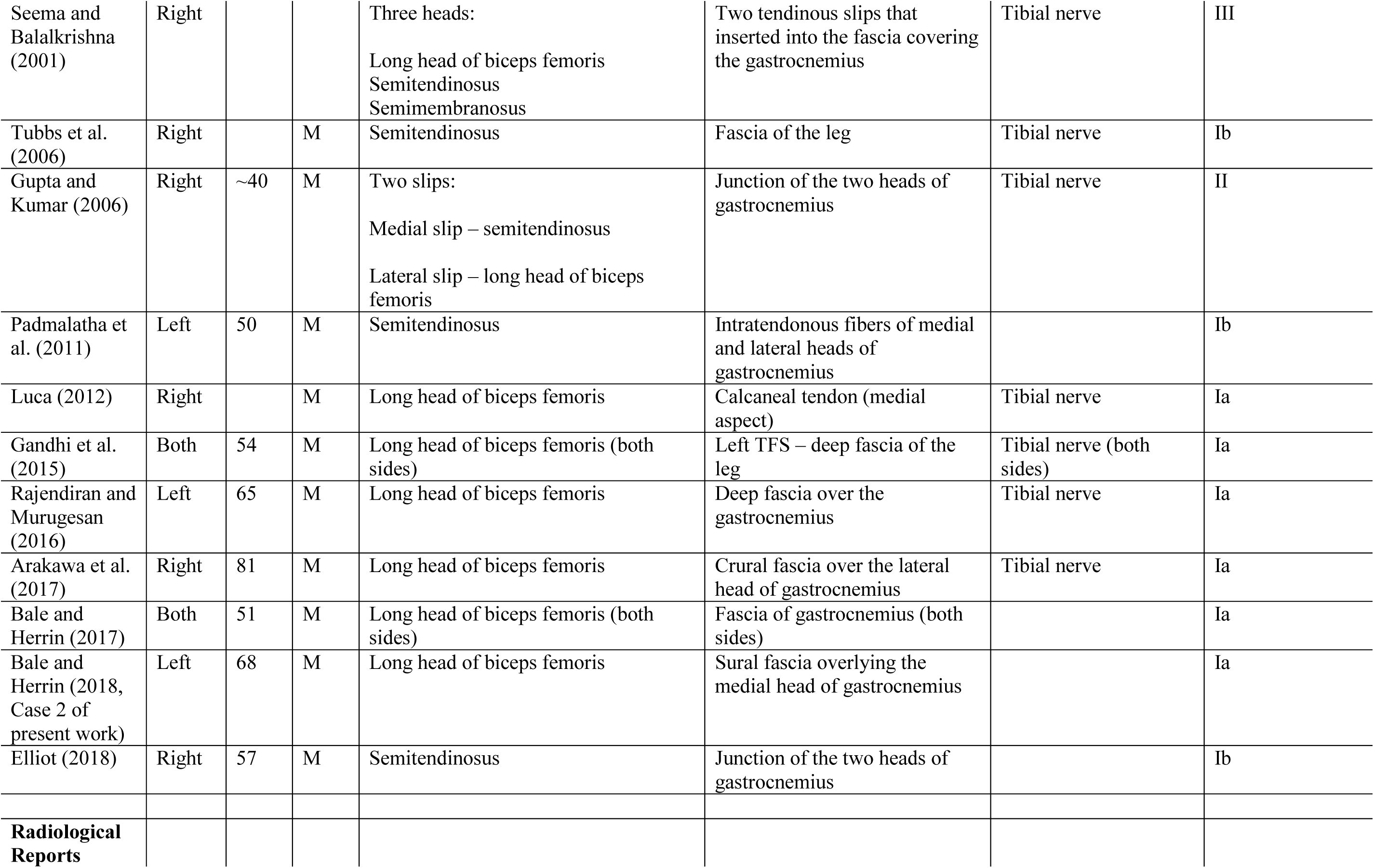
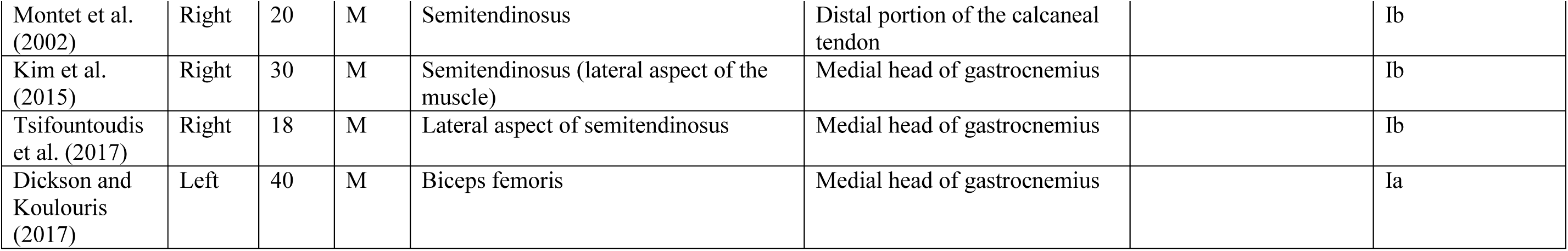
Summary of cadaveric and radiological case reports of TFS.

## DISCUSSION

Tensor fasciae suralis is considered a rare muscle. We report its prevalence to be 1.3% based on dissection of 236 lower limbs. In a report of a case of TFS^8^, the authors noted that the muscle was not seen in a study of 300 cadaveric limbs in which the popliteal fossae were dissected to identify muscular and neurovascular variations^30^. To the best of our knowledge the present work is the first study dedicated to determining the prevalence of TFS.

Based on the literature, we propose a classification system for the subtypes of TFS. The proximal attachments of TFS and number of heads present were used as defining criteria to differentiate between subtypes. With few exceptions, the insertion of TFS (regardless of subtype) was the deep fascia of the leg, gastrocnemius, or calcaneal tendon. If adapted, this naming convention could help dispel misconceptions that are associated with TFS. For instance, TFS has been said to arise most commonly from the semitendinosus muscle^13,31,32^ or solely from semitendinosus without mention of the muscle’s other possible origins^12,27,33^. Based on the literature such an origin from semitendinosus is slightly less common than an origin from the long head of biceps femoris, and therefore we suggest that these subtypes be termed Ib and Ia, respectively. A second misconception related to TFS revolves around the definition of the muscle itself. The name “tensor fasciae suralis (ischioaponeuroticus) has been given to a muscular slip passing from one or other of the hamstring muscles to the fascia of the back of the leg”^34^. Interestingly, this description of TFS was included within a portion of text that was dedicated to variations of the semimembranosus muscle. Quain’s definition of TFS was referenced by Barry and Bothroyd^23^ in their seminal report of the muscle variant, which appears to have cemented the notion that TFS can arise from any hamstring muscle. However, none of the reports summarized in this work describe the semimembranosus as the sole proximal attachment of TFS. Only one case involves semimembranosus at all; an unusual three-headed TFS case where one of the heads took origin from semimembranosus^9^. To make matters more confusing, a case of an “accessory semimembranosus” muscle (which arose from the semimembranosus, coursed through the popliteal fossa, and inserted at the proximal portion of the medial head of the gastrocnemius) has been reported^35^. One could argue that this muscle was tensor fasciae suralis instead of accessory semimembranosus. The long-standing notion that TFS can arise from any hamstring muscle does not seem to be reinforced by the literature, and as such we do not include within our classification system a distinction for a subtype of TFS that arises solely from semimembranosus.

The way TFS emerges and persists during development is not clear. It has been suggested that TFS represents a muscular persistence of embryological fibrous connections that span the tendon of biceps femoris and the deep fascia of the leg^10^. A particularly interesting case of identical twin fetuses was reported by Mogi^25^: TFS was present in the right popliteal fossa of one twin and a third head of the gastrocnemius was present in the right popliteal fossa of the other twin. This rare case may suggest that TFS and the third head of gastrocnemius have a similar developmental origin. The manner in which TFS attaches to the crural fascia is similar to muscles found in some mammals^1,22^.

Tensor fasciae suralis is not strictly of interest to anatomists as it does have clinical implications in certain instances. The muscle may present as a painless mass of the popliteal fossa^27,33^, which could lead to unexpected findings during physical examination^32,36^. While rare, athletic injuries of TFS have been reported. It is speculated that TFS may cause compressive injuries of the popliteal vein^8^ and the sciatic, tibial, and sural nerves^13^. With respect to Case 2 of the present work, the position of the distal aspect of TFS relative to the median sural nerve suggests that this nerve could potentially be compressed by TFS.

Tensor fasciae suralis is a rare supernumerary muscle of the posterior thigh and popliteal fossa. We report three TFS muscles that were identified in a study of 236 formalin-fixed cadaveric lower limbs (prevalence = 1.3%). Based on the literature, we suggest a classification system to organize the subtypes of TFS.

## Data Availability

All data used for the manuscript is included in the manuscript.

## ACKNOWLEDGEMENTS

The authors wish to thank individuals who donate their bodies and tissues for the advancement of education and research. They also thank Bill Borman, Michael Carnes, Jayme Gallegos, Katie Lockwood, and the students of University of Western States.

## Notable variants of the popliteal fossa are briefly discussed below for comparison with tensor fasciae suralis

### Third head of the gastrocnemius (gastrocnemius tertius, caput tertium of Kelch)

The third head of the gastrocnemius is the most common variation of the gastrocnemius muscle. The third head may be large and bulky, or it may be tendinous and inconspicuous. The reported prevalence of the third head of the gastrocnemius has varied from 1.9%^1^ to 5.5%^2^. A range of possibilities exist with respect to the accessory head’s proximal attachment(s), distal attachment(s), and course relative to the popliteal neurovascular bundle. Twelve variations of the third head of the gastrocnemius have been reported^3^.

### Accessory plantaris (bicipital plantaris of Hall)

The accessory plantaris muscle belly may arise from a range of sites and typically fuses with the muscle belly of plantaris^4–6^. In a retrospective review of 1000 consecutive MRI exams of patients with chronic or acute knee issues, the accessory plantaris muscle was identified in 63 patients (6.3%)^4^. In a recent cadaveric study, the accessory plantaris muscle was found in 3 of 50 limbs (6%)^5^.

### Accessory popliteus (of Fabrice, popliteus biceps)

Accessory popliteus is a rare variant that exhibits a range of potential origins (lateral femoral condyle, lateral head of gastrocnemius, capsule of the knee joint, or posterior cruciate ligament) and inserts near the soleal line on the medial portion of the tibia^6^. According to Bartoníček, Gruber identified an accessory popliteus in 11 of 695 dissected limbs (1.6%)^7^. An MRI report of what was likely accessory popliteus has been published^8^.

### Transverse muscle of popliteal fossa

Several authors have documented an anomalous muscle that passes transversely through the popliteal fossa, spanning the tendon of biceps femoris and the medial head of the gastrocnemius ^9–11^. Despite the similarities between these cases, the innervation of the muscle has been found to be from the common fibular nerve^9,10^, tibial nerve^11^, or lateral sural cutaneous nerve^12^. This transverse muscle of the popliteal fossa lacks an agreed upon name. It has been speculated to be a conversion of fascia lata into muscle^9^, a derivation from the short head of biceps femoris^10^, or a variation of the third head of the gastrocnemius^13^. Although similar in its attachments, this transverse muscle is fundamentally different than TFS as the latter passes longitudinally through the popliteal fossa and not transversely like the former.

## REFERENCES

1. Bergman R, Thompson S, Afifi A. Catalog of Human Variation. Balitmore, Munich: Urban & Schwarzenberg; 1984.

2. Tubbs RS, Salter EG, Oakes WJ. Dissection of a rare accessory muscle of the leg: the tensor fasciae suralis muscle. Clin Anat. 2006;19(6):571–572.

3. Schaeffer JP. On two muscle anomalies of the lower extremity. Anat Rec. 1913;7(1):1–7.

4. Kelch W. Abweichung des biceps femoris. Beiträge z pathol Anat. 1813;Bd. 8:art 36,42.

5. Nonaka T, Ishii S. A case of m. tensor fasiae suralis. J Correct Med. 1954;2:25–27.

6. Miyauchi. A case of tensor fasciae suralis muscle. Acta Med Nagasaki. 1985;30(1-3):285–288.

7. Sinav A, Gümüşalan Y, Arifoğlu Y, Onderoğlu S. Accessory muscular bundles arising from biceps femoris muscle. Kaibogaku Zasshi. 1995;70(3):245–247.

8. Somayaji SN, Vincent R, Bairy KL. An anomalous muscle in the region of the popliteal fossa: case report. J Anat. 1998;192(Pt 2):307–308.

9. Seema S, Balalkrishna. Tensor fascia suralis - a case report. In: 49th Annual Conference of the Anatomical Society of India. Karnataka, India; 2001.

10. Gupta R, Kumar BSS. An anomalous muscle in the region of the popliteal fossa: A case report. J Anat Soc India. 2006;55(2):65–68.

11. Luca C. The tensor fasciae suralis muscle - case report. Rev Rom Anat Funct si Clin Macro-si Microsc si Antropol. 2009;8(1):501–503.

12. Padmalatha K, Prakash B, Mamatha Y, Ramesh B. Ischioaponeuroticus / tensor fascia suralis. Int J Anat Var. 2011;4(1):104–105.

13. Gandhi KR, Wabale RN, Farooqui MS. Bilateral presentation of tensor fascia suralis muscle in a male cadaver. Int J Anat Res. 2015;3(4):1745–1748.

14. Rajendiran R, Murugesan A. Unilateral tensor fascia suralis: a case report. Brunei Darussalam J Heal. 2016;6(2):94–98.

15. Turner M. Report on the progress of anatomy (M. tensor fasciae suralis and M. tensor fasciae poplitealis). J Anat Phsiol. 1872;6:442.

16. Arakawa T, Kondo T, Tsutsumi M, Watanabe Y, Terashima T, Miki A. Multiple muscular variations including tenuissimus and tensor fasciae suralis muscles in the posterior thigh of a human case. Anat Sci Int. 2017;92(4):581–584.

17. Bale LSW, Herrin SO. Bilateral tensor fasciae suralis muscles in a cadaver with unilateral accessory flexor digitorum longus muscle. Case Rep Med. 2017;2017:1–4.

18. Elliott GE. A muscle variant in the region of the popliteal fossa associated with the semitendinosus muscle. Int J Anat Var. 2018;11(4):127–128.

19. Gruber W. Ûber eine variante des vom musculus semitendinosus abgehenden musculus tensor fasciae suralis. Bull l’Acad Imp des Sci St Petersbg. 1873;18:184–186.

20. Gruber W. Über die ungewöhnlichen musculi tensores fasciae suralis beim menschen. Bull Acad Imp Sci Saint-pétersbg. 1879;25:230–236.

21. Halliburton W. Remarkable abnormality of the musculus biceps flexor cruris. J Anat Physiol. 1881;15:296–299.

22. Klaatsch H. Über eine dem Tenuissimusa ähnliche Variation am Biceps femoris des Menschen. Anat Anz. 1911;38:305.

23. Barry D, Bothroyd JS. Tensor fasciae suralis. J Anat. 1924;58(Pt 4):382-383.

24. Kawai M. Über drei muskelanomalien. Acta Anat Nippon. 1935;8:270–275.

25. Mogi E. Muskelvarietaten der unteren Extremitaten bei den japanischen Zwillingsfeten. Okajimas Folia Anat Jpn. 1940;19(1):93–95.

26. Mudiraj N, Dhobale M, Joshi U. Tensor fasciae suralis - an unusual variation. Biomirror. 2012;3(1):5–7.

27. Chason DP, Schultz SM, Fleckenstein JL. Tensor fasciae suralis: Depiction on MR images. Am J Roentgenol. 1995;165(5):1220–1221.

28. Olave E, Prates JC, Gabrielli C, Pardi P. Morphometric studies of the muscular branch of the median nerve. J Anat. 1996;189(Pt 2):445–449.

29. George BM, Nayak SB, Marpalli S. Clinical importance of tensor fasciae suralis arising from linea aspera along with short head of biceps femoris: A rare anomaly. Anat Cell Biol. 2019;52(1):90–92. doi:10.5115/acb.2019.52.1.90

30. Somayaji S, Nayak S, Bairy K. Recorded anomalies of the popliteal artery. J Anat Soc India. 1996;45:23–26.

31. Kim KH, Shim JC, Lee GJ, Lee KE, Kim HK, Suh JH. MR Imaging and ultrasonographic findings of tensor fasciae suralis muscle : A case report. J Korean Soc Radiol. 2015;73(4):249–251.

32. Dickson T, Koulouris G. Acute posterior thigh pain in an athlete. Skeletal Radiol. 2017;46(1):141–142.

33. Montet X, Sandoz A, Mauget D, Martinoli C, Bianchi S. Sonographic and MRI appearance of tensor fasciae suralis muscle, an uncommon cause of popliteal swelling. Skeletal Radiol. 2002;31(9):536–538.

34. Quain J. Quain’s Elements of Anatomy. Volume II - Part II. 10th ed. London, New York: Longmans, Grenn, & Co; 1894.

35. Stoane JM, Gordon DH. MRI of an accessory semimembranosus muscle. J Comput Assist Tomogr. 1995;19(1):161–162.

36. Tsifountoudis I, Kalaitzoglou I, Papacostas E, Malliaropoulos N. Tensor fasciae suralis muscle. Clin J Sport Med. July 2017:1.

## References

1. Koplas MC, Grooff P, Piraino D, Recht M. Third head of the gastrocnemius: an MR imaging study based on 1,039 consecutive knee examinations. Skeletal Radiol. 2009;38(4):349–354.

2. Tochiara J, Onosawa T. M. gastrocnemius tertius in Japanese. Acta Anat Nippon. 1932;5:589–600.

3. Bergman RA, Walker CW, EI-Khour GY. The third head of gastrocnemius in CT images. Ann Anat. 1995;177(3):291–294.

4. Herzog RJ. Accessory Plantaris Muscle: Anatomy and Prevalence. HSS J. 2011;7(1):52–56.

5. Jayasree K, Ashalatha P, Nair S,. Variations of muscle plantaris: anatomical and clinical implications. J Evol Med Dent Sci. 2016;5(45):2892–2896.

6. Tubbs RS, Shoja MM, Loukas M. Bergman’s Comprehensive Encyclopedia of Human Anatomic Variation. Wiley-Blackwell; 2016.

7. Bartoníček J. Rare bilateral variation of the popliteus muscle. Anatomical case report and review of the literature. Surg Radiol Anat. 2005;27(4):347–350.

8. Duc SR, Wentz KU, Käch KP, Zollikofer CL. First report of an accessory popliteal muscle: Detection with MRI. Skeletal Radiol. 2004;33(9):550.

9. Schaeffer JP. On two muscle anomalies of the lower extremity. Anat Rec. 1913;7(1):1–7.

10. Okamoto. An anomalous muscle in the superficial region of the popliteal fossa, with special reference to its innervation and derivation. Ann Anat. 2004;186:555–559.

11. Kim BS, Kim SH, Cho SS, Yoon SP. A rare muscular variation in the superficial region of the popliteal fossa. Surg Radiol Anat. 2014;36(7):721–723.

12. Kim D-I, Kim H-J, Shin C, Lee K-S. An abnormal muscle in the superficial region of the popliteal fossa. Anat Sci Int. 2009;84(1-2):61–63.

13. Guo PJ, Kim J, Rubin R. How video production affects student engagement: An empirical study of MOOC videos. In: Proceedings of the 1st ACM Conference on Learning at Scale.

